# An improved methodology for estimating the prevalence of SARS-CoV-2

**DOI:** 10.1101/2020.08.04.20168187

**Authors:** Virag Patel, Catherine McCarthy, Rachel A Taylor, Ruth Moir, Louise A Kelly, Emma L Snary

## Abstract

Since the identification of Coronavirus disease 2019 (COVID-19) caused by severe acute respiratory syndrome coronavirus 2 (SARS-CoV-2) in China in December 2019, there have been more than 17 million cases of the disease in 216 countries worldwide. Comparisons of prevalence estimates between different communities can inform policy decisions regarding safe travel between countries, help to assess when to implement (or remove) disease control measures and identify the risk of over-burdening healthcare providers. Estimating the true prevalence can, however, be challenging because officially reported figures are likely to be significant underestimates of the true burden of COVID-19 within a community. Previous methods for estimating the prevalence fail to incorporate differences between populations (such as younger populations having higher rates of asymptomatic cases) and so comparisons between, for example, countries, can be misleading. Here, we present an improved methodology for estimating COVID-19 prevalence. We take the reported number of cases and deaths (together with population size) as raw prevalence for the population. We then apply an age-adjustment to this which allows the age-distribution of that population to influence the case-fatality rate and the proportion of asymptomatic cases. Finally, we calculate the likely underreporting factor for the population and use this to adjust our prevalence estimate further. We use our method to estimate the prevalence for 166 countries (or the states of the United States of America, hereafter referred to as US state) where sufficient data were available. Our estimates show that as of the 30^th^ July 2020, the top three countries with the highest estimated prevalence are Brazil (1.26%, 95% CI: 0.96 – 1.37), Kyrgyzstan (1.10%, 95% CI: 0.82 – 1.19) and Suriname (0.58%, 95% CI: 0.44 – 0.63). Brazil is predicted to have the largest proportion of all the current global cases (30.41%, 95%CI: 27.52 – 30.84), followed by the USA (14.52%, 95%CI: 14.26 – 16.34) and India (11.23%, 95%CI: 11.11 – 11.24). Amongst the US states, the highest prevalence is predicted to be in Louisiana (1.07%, 95% CI: 1.02 – 1.12), Florida (0.90%, 95% CI: 0.86 – 0.94) and Mississippi (0.77%, 95% CI: 0.74 – 0.81) whereas amongst European countries, the highest prevalence is predicted to be in Montenegro (0.47%, 95% CI: 0.42 - 0.50), Kosovo (0.35%, 95% CI: 0.29 - 0.37) and Moldova (0.28%, 95% CI: 0.23 - 0.30). Our results suggest that Kyrgyzstan (0.04 tests per predicted case), Brazil (0.04 tests per predicted case) and Suriname (0.29 tests per predicted case) have the highest underreporting out of the countries in the top 25 prevalence. In comparison, Israel (34.19 tests per predicted case), Bahrain (19.82 per predicted case) and Palestine (9.81 tests per predicted case) have the least underreporting. The results of this study may be used to understand the risk between different geographical areas and highlight regions where the prevalence of COVID-19 is increasing most rapidly. The method described is quick and easy to implement. Prevalence estimates should be updated on a regular basis to allow for rapid fluctuations in disease patterns.

## Introduction

The ability to accurately estimate (sub)-population prevalence is a fundamental pillar of effective infectious disease management, allowing communities to step up (or down) resources when needed. Comparisons of prevalence estimates between different communities can inform policy decisions regarding safe travel between communities (including international travel between different countries). Understanding differences between communities and within the same community at different time points, can help to identify any lessons learnt and help to guide preparedness for the next epidemic (and even different stages of the same epidemic). Moreover, the ability to understand the burden on healthcare providers in real-time, together with an accurate understanding of healthcare capacity, is vitally important in understanding whether demand might outstretch resources and identifying at-risk communities.

Since the identification of Coronavirus disease 2019 (COVID-19) caused by severe acute respiratory syndrome coronavirus 2 (SARS-CoV-2) in China in December 2019, there have been more than 17.6 million reported cases of the disease in 216 countries worldwide (data correct as of 2^nd^ August 2020 (WHO, 2020)). Officially reported figures of the number of COVID-19 cases are likely to be significant underestimates of the true number within a community, due to the high levels of asymptomatic carriage or mild cases which may not be diagnosed. Official figures are also estimates of total incidence (number of confirmed cases) rather than prevalence (number of active cases at one point in time), and therefore cannot reliably assess the burden of a disease within a community. Moreover, there are likely to be significant country (or even community) level differences in underreporting rates. Logistical barriers, such as sourcing test kits and implementing testing schemes can limit access to testing. It has been suggested that Asia-Pacific regions, which have incorporated lessons learnt from past epidemics such as H5N1 and SARS, have significantly stronger pandemic preparedness plans than many European countries, allowing them to step up testing rapidly (Coker and Mounier-Jack, 2006). Moreover, personal barriers such as concerns regarding financial consequences of a positive test, societal stigma and remote communities can decrease an individual’s willingness to get tested. This was perceived as a particular barrier during the Ebola outbreak in West Africa from 2013-2016 (Shultz *et al*., 2016). It is clear that accurate estimates of disease prevalence must not rely solely on reported cases or deaths, but must also incorporate differences in under-reporting rates.

Different models have been developed to try to recreate and forecast incidence rates of COVID-19 cases across the world, including time series models (Ceylan, 2020), artificial intelligence based approaches (Al-Qaness *et al*., 2020; Saba and Elsheikh, 2020) and publically available data including Google searches (Husnayain, Fuad and Su, 2020). These methods are useful for fitting to current data and forecasting future trends, but represent incidence rates which therefore fail to help in understanding disease burden within a community. Additionally, they do not provide methods to understand subtle differences between communities, such as access to testing, differences in case-fatality rates (CFR) and any consequences of heterogeneous age distributions. Other methods of estimating COVID-19 prevalence rates have assumed a single value for CFR (Russell *et al*., 2020). However, studies have shown that both CFR (Riou *et al*., 2020; Verity *et al*., 2020) and asymptomatic rates (Davies *et al*., 2020) have varied by age, with elderly subgroups being at higher risk of death if infected but a lower likelihood of being asymptomatic. Therefore, accurately estimating the number of COVID-19 cases based on the age distribution within the population is important. We aim to produce an improved methodology for estimating prevalence rates of COVID-19 and use this to estimate country-level prevalence, and state level for the United States of America (USA). This model incorporates heterogeneity in age populations by using age-specific CFRs and asymptomatic rates. Moreover, the method allows us to assess differences in under-reporting rates and highlight countries where the barriers to testing are highest. The results of this study may be used to understand risk between different geographical areas and, if updated regularly, highlight regions where the prevalence of COVID-19 is increasing most rapidly.

## Methods

The key to estimating the prevalence of SARS-CoV-2 in each country or state is the estimation of the true number of cases in each country or state. We use a two-step approach to estimate this number. The first is to account for a country’s Case to Fatality Ratio (CFR) and the proportion of cases that may be asymptomatic. Both of these adjustments are age-standardised calculations to allow for heterogeneity in age populations. The second step is to recalibrate the estimated expected number of cases by adjusting for the number of tests a country or state has performed.

### Data

Data on the numbers of cases and deaths and number of tests for 209 countries and territories were collated and provided by Public Health England (PHE) on the 30^th^ of July 2020, based on either country specific public updates or data collated by WHO (WHO, 2020). The dataset includes the daily and total numbers of COVID-19 cases and formally notified COVID-19 deaths but may be subject to some variation due to differences in countries case definitions. Countries or territories with either no test data (n = 14), no age distribution data (n = 11) or no recorded deaths (n = 18) were excluded from the dataset. Population estimates for the remaining 166 countries and territories were extracted from the Our World in Data website (Ritchie, 2020). Data for US states were also provided by PHE, collected from the Covidtracking.com website (Atlantic, 2020) of the same date. Only one US territory, American Samoa, was excluded as there were no recorded cases.

Some countries and states have been retrospectively reducing their official numbers of cases and deaths, leading to negative numbers of new cases and deaths on certain days. In order to prevent biased prevalence estimates, the daily numbers of deaths or cases with negative values were replaced by taking the average between the reported numbers for the closest dates before and after the date that report a non-negative number. For example if a country retrospectively reduced the number of cases on the 25^th^ July, the number of estimated cases for this date would be the average number of reported cases between 24^th^ June and 26^th^ July assuming they were both non-negative. If the 26^th^ of July also recoded a negative daily number then the number for the 27^th^ July would be used and so on. For countries that have retrospectively reduced their official reported cases or deaths on the 30^th^ July, it was assumed to be the same as the 29^th^ July. 13 countries have reported the number of people tested rather than the total number of tests performed. For these countries, we assumed that the number of tests was the same as the number of people tested.

Data on age composition for each country and territory was extracted from the United Nations Department of Economic and Social Affairs website (UN, 2020). For US states, age distribution data was extracted from the US census website (USCB, 2020).

### Prevalence estimation

The prevalence for each country (*c*) was calculated as:

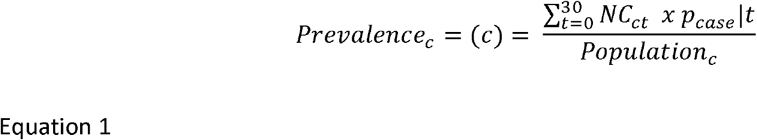

where *t* is the *t^th^* day of the disease period, *c* denotes the *c^th^* country, *NC_ct_* is the number of new cases for the *c^th^* country on the *t^th^* day and *p_case_*|*t* is the probability a confirmed case is still in their disease period (i.e. still a case) at day *t*. The probability of being a case was dependent upon the number of days since exposure to SARS-CoV-2. We simulated a population of one million infected individuals and estimated their incubation period, duration of symptom presentation and duration of infectivity based on published disease progression distributions (Davies, Kucharski, *et al*., 2020). The final day that each case was within their disease period (defined as being either infectious or having symptoms) was recorded. Due to the potentially very long tail of the distribution of disease period (primarily those with very severe disease), this analysis only considers individuals who are within the first 30 days of infection; any individuals with a disease period of over 30 days were assumed to be disease free at day 30 instead. **Error! Reference source not found**, shows the distribution for the probability that a confirmed case is still classed as a case at day *t* (*p_case_*|*_t_*). This is calculated by:

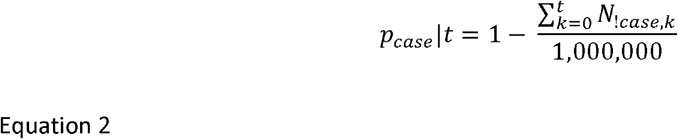

where *k =* the *k^th^* day such that 0 ≤ *k* ≤ *t, N*_!_*_case_* is the number of simulated individuals that are no longer within their disease period (classed as a case).

To calculate the number of new cases per day for each country (*NC_ct_*, **Error! Reference source not found.)**, we first estimate the total number of cases after adjusting for country-specific Case to Fatality Ratio (CFR_c_) and asymptomatic rates (ASR_C_) for each country. Let us denote this as *E*(*TC|CFR.ASR*). The definition of an asymptomatic case is the same as that defined as a subclinical case by (Davies, Klepac, *et al*., 2020). “Asymptomatic” that have no symptoms at all, and “paucisymptomatic” is sometimes used for those with very mild symptoms that may not be noticed or reported, even though they occur.”

The CFR is used to predict the number of new cases based on the reported number of deaths. The CFR for each country (*CFR_c_*) is calculated by:

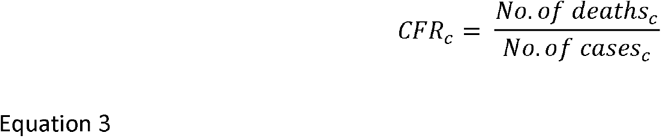

We assume that:

1. *CFR_c_* is constant across all countries.
2. *ASR_c_* is constant across all countries.
3. The number of reported deaths is correct.

Providing these assumptions hold, the expected number of cases per country can be estimated by:

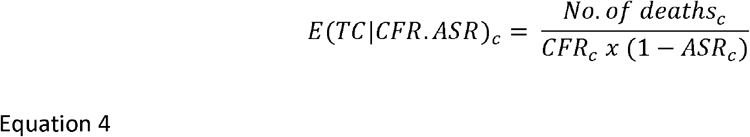

Studies by Verity et al. (2020) and Riou, Hauser, Counotte, and Althaus (2020) found that the CFR varied across age groups with elderly subgroups being at higher risk of death if infected. Both studies produced similar CFR estimates for varying age ranges, however the Verity study reported smaller confidence intervals and therefore these estimates were used for these calculations. Similarly, Davies, Klepac, et al. (2020) found that the ASR also varied by age groups. As both the CFR and ASR estimates are obtained for studies in China, the CFR and ASR for other countries may differ significantly due to differences in age distributions. Therefore age distribution data combined with these age categorised CFR and ASR estimates are used to estimate country specific *CFR_c_* and *ASR_c_*. For each country, we calculate *E*(*TC|CFR. ASR*)*_c_* as:

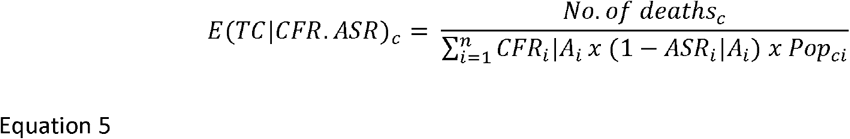

where, *CFR_i_|A_i_* is the CFR for the corresponding *f^h^* age group (*A_i_*)*, n* is the total number of age groups. *ASR_i_|A_i_* is the ASR for the corresponding *i^th^* age group (*A_i_*)*. Pop_ci_* is the proportion of the population in the *i^th^* age group within the *c^th^* country.

The second step is to recalibrate *E*(*TC|CFR. ASR*)*_c_* such that it takes into account the amount of testing performed by each country. The *E*(*TC|CFR.ASR*)*_c_* estimate is likely to be an overestimate, especially in countries with a relatively high number of deaths. For example, in the UK, *E*(*TC|CFR.ASR*)*_UK_* = 3,004,492 (2,903,113, 3,149,334) which is 4.55% of the country’s population, similarly in Belgium the *E*(*TC|CFR. ASR*)*_Bel_* estimated that 5.37% of the country’s population may have contracted SARS-CoV-2. The main reason for this overestimation is because we have assumed that there has been no response from the country to the outbreak such as testing, implementing lockdown and quarantine procedures. The *CFR_i_|A_i_* estimates that we have used, are based on aggregated data obtained from the earliest points of the outbreak in China between 1^st^ January (Verity et al. 2020). The first city to lockdown was Wuhan on 23^rd^ January 2020.

One way to recalibrate the predicted number of cases is to use the data on the number of tests performed by each country. It is assumed a country that has done more testing per population has more reliability in their reporting of the number of cases. This may also give an indication of a country’s response to the outbreak. A country that that has rigorously tested its population is likely to be responding in a positive way to the outbreak compared to a country that isn’t.

For this calibration step, the 166 countries with data for the number of tests or number of people tested was used. We assume that the expected number of cases, calculated by adjusting for *CFR_c_* and *ASR_C_*, (*E*(*TC|CFR.ASR*)*_c_*) is an overestimate for these countries. It is also likely that the reported number of cases for each country (let us call this *R*(*TC*)*_c_*) is an underestimate, therefore a natural range between these values can be defined and the “true” expected number of cases (let us call this 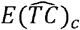) is likely to lie within this range. Therefore we apply following linear model:

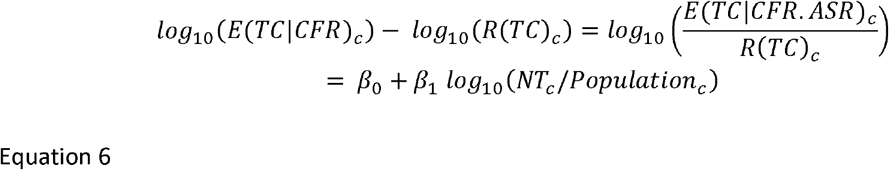

where, 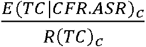 is an underestimation factor of the CFR estimate compared to the reported number and *NT_c_* is the number of tests performed by the c^th^ country. The regression coefficient based on the data from 166 countries was *β*_1_ = −0.2901 (p =7.36×10^−15^) and *β*_0_ = 0.019. This gives a good indication that the number of tests will explain some of the variation between the expected and reported number of tests (Figure 1). Equation 6 can then be rearranged with the estimated regression coefficients to predict the adjusted expected number of cases (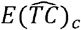).

**Figure 1:**
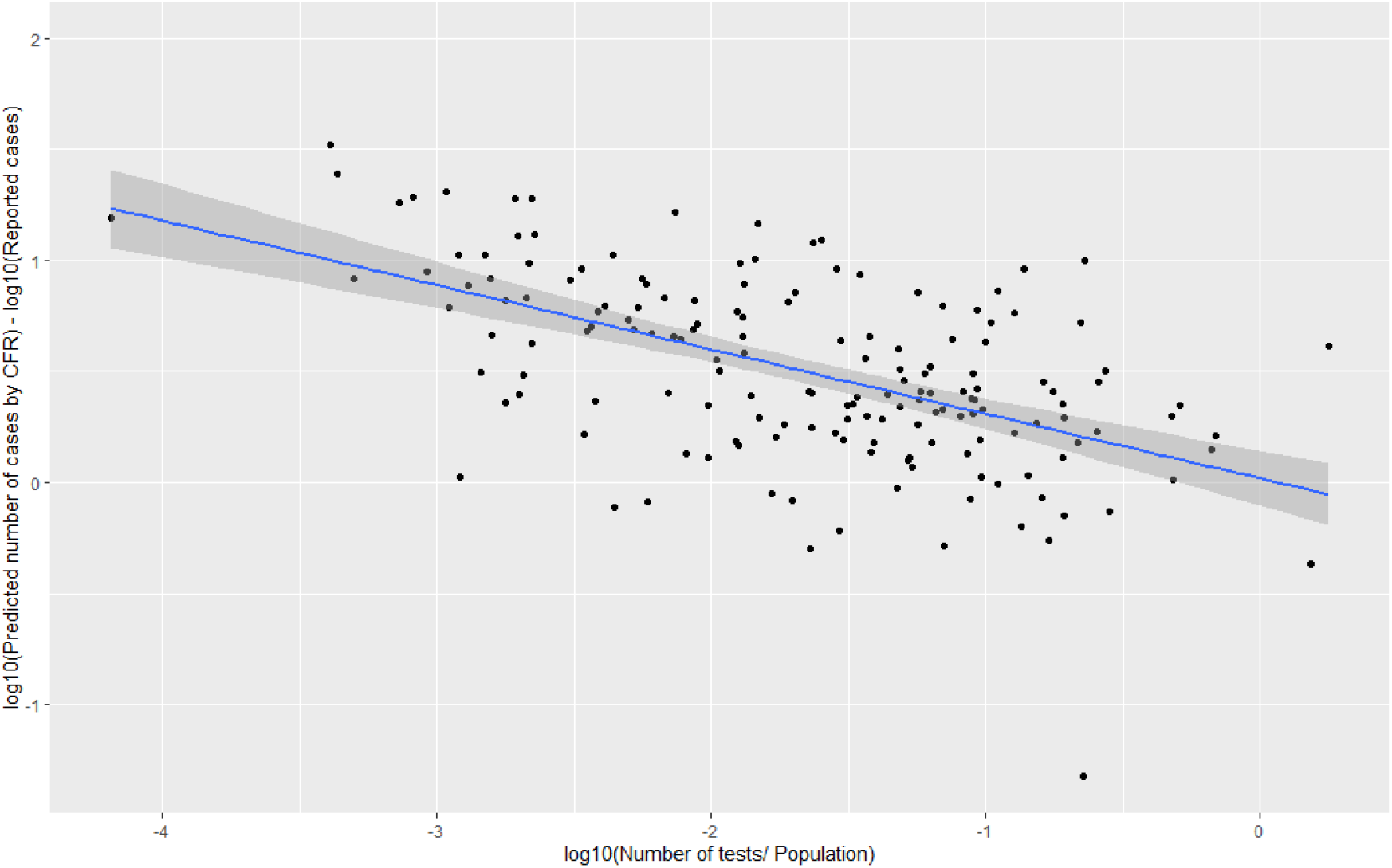
Scatterplot showing the relationship between the number of tests per population and the CFR estimated underreporting factor from 166 countries.

This re-calibration step provides a simple linear adjustment based on the number of tests per population. The more testing performed by a country, the closer the number of predicted cases estimate 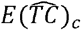 will be to the number of reported cases *R*(*TC*)*_c_*. It also puts less emphasis on the number of deaths assumption in the CFR calculation, which may be biased due to a reporting delay and under-reporting.

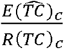 is the estimated under-reporting factor after re-calibrating for the number of tests a country has performed, which can be multiplied by the number of reported cases over the 30 day period to obtain the predicted number of new cases. We apply the recalibration step for all 166 countries and the 51 US states and territories separately as there may be some within country heterogeneity to consider.

## Results

Our results show that the top five countries with the highest estimated prevalence (as of 30th July 2020) are Brazil (1.26%, 95% CI: 0.96 – 1.37), Kyrgyzstan (1.10%, 95% CI: 0.82 – 1.19), Suriname (0.58%, 95% CI: 0.44 – 0.63), Panama (0.58%, 95% CI: 0.52 – 0.62) and Montenegro (0.47%, 95% CI: 0.42 – 0.50) (Figure 2). Brazil is predicted to have the largest proportion of all the current global cases (30.41%, 95% CI: 27.52 – 30.84), followed by the USA (14.52%, 95% CI: 14.26 – 16.34), India (11.23%, 95% CI: 11.11 – 11.24), Mexico (3.43%, 95% CI: 3.29 – 3.44) and Russia (2.73%, 95% CI: 2.68 – 3.08).

**Figure 2:**
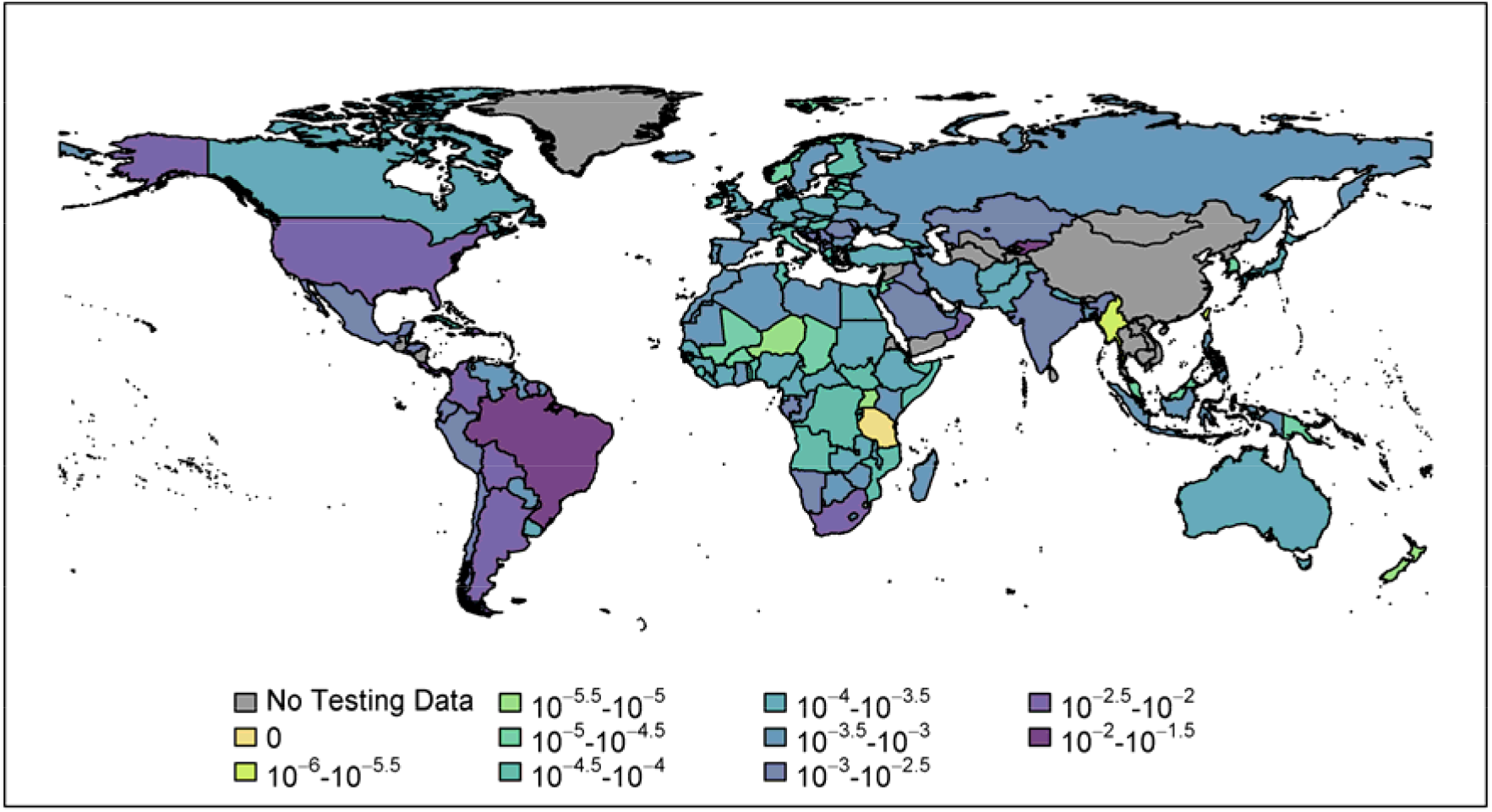
International prevalence of COVID-19 as of 30^th^ July 2020. Colours show predicted prevalence rates which have been adjusted for reported cases and deaths given country-level differences in age-distributions and underreporting rates. Colours range from light green (lowest prevalence) to deep purple (highest prevalence).

Amongst the US states, the highest prevalence is predicted to be in Louisiana (1.07%, 95% CI: 1.02 - 1.12), Florida (0.90%, 95% CI: 0.86 - 0.94), Mississippi (0.77%, 95% CI: 0.74 - 0.81), Tennessee (0.71%, 95% CI: 0.68 - 0.75) and Georgia (0.60%, 95% CI: 0.57 - 0.62) (Figure 3). Interestingly, we found that the raw prevalence estimates for these states where higher than the CFR and ASR adjusted estimates, suggesting that the number of cases is much greater than predicted for these particular states. Amongst European countries, the highest prevalence countries are mostly situated in southeast Europe: Montenegro (0.47%, 95% CI: 0.42 - 0.50), Kosovo (0.35%, 95% CI: 0.29 - 0.37), Moldova (0.28%, 95% CI: 0.23 - 0.30), Luxembourg (0.18%, 95% CI: 0.18 - 0.19%) and North Macedonia (0.18%, 95% CI: 0.16 - 0.19%). Out of the countries in mainland Europe, the prevalence values were highest in Luxembourg (0.18%, 95% CI: 0.18 - 0.19%), Spain (0.08%, 95% CI: 0.08 - 0.09%), Monaco (0.07%, 95% CI: 0.06 - 0.07%), Belgium (0.05%, 95% CI: 0.05 - 0.06%) and Sweden (0.05%, 95% CI: 0.05 - 0.06%).

**Figure 3:**
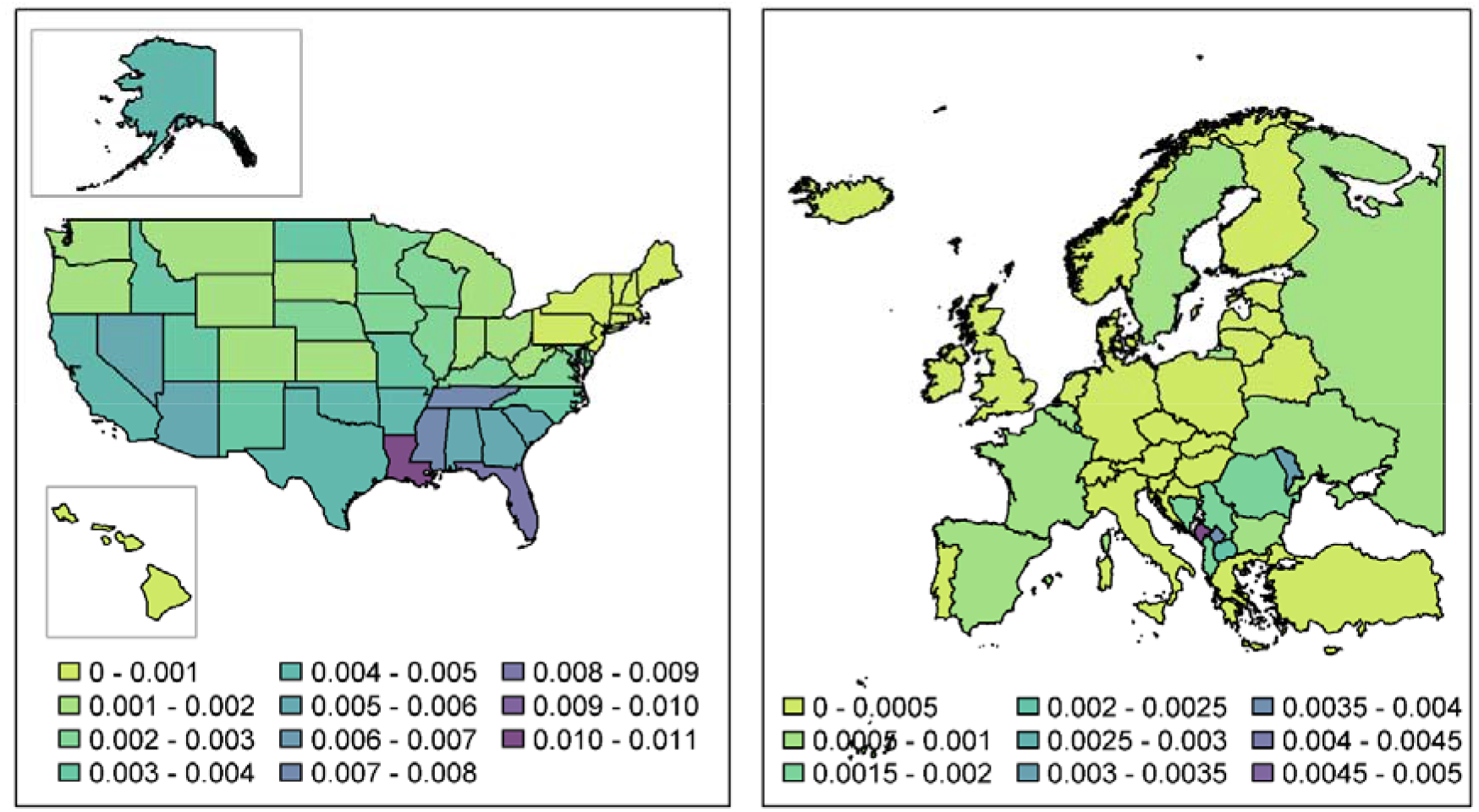
Prevalence of COVID-19 per US state (left figure) and European country (right figure) as of 30^th^ July 2020. Colours show predicted prevalence rates which have been adjusted for reported cases and deaths given country-level differences in age-distributions and underreporting rates. Colours range from light green (lowest prevalence) to deep purple (highest prevalence).

Our method for calculating the prevalence has two fundamental aspects. The raw prevalence estimate (based on reported cases and deaths), is first adjusted to allow for differences in age distributions between countries (thereby affecting both CFR and asymptomatic rates). Secondly, an underreporting factor is applied in order to calculate the final predicted prevalence (Figure 4). According to the countries which are estimated to have the 25 highest prevalence estimates, using the raw prevalence alone would rank Bahrain as highest (0.24%), followed by Panama (0.23%) and Oman (0.23%). However, differences in age distributions between countries means that country comparisons cannot be made without first adjusting for differences in CFR and ASR. Countries with younger populations (or possibly enhanced healthcare provisions) would see a large difference between the raw prevalence and age-adjusted prevalence as many cases would not result in many fatalities (such as Kyrgyzstan, Sint Maarten and Peru in our analysis). Comparing the age adjusted prevalence with the final predicted prevalence allows us to compare the underreporting factor between countries. In our analysis, Israel (34.19 tests per predicted case), Bahrain (19.82 tests per predicted case) and Palestine (9.81 tests per predicted case) test a higher proportion of their cases than other countries. In comparison, Kyrgyzstan (0.04 tests per predicted case), Brazil (0.04 tests per predicted case) and Suriname (0.29 tests per predicted case) test a lower proportion, suggesting many cases remain undetected.

**Figure 4:**
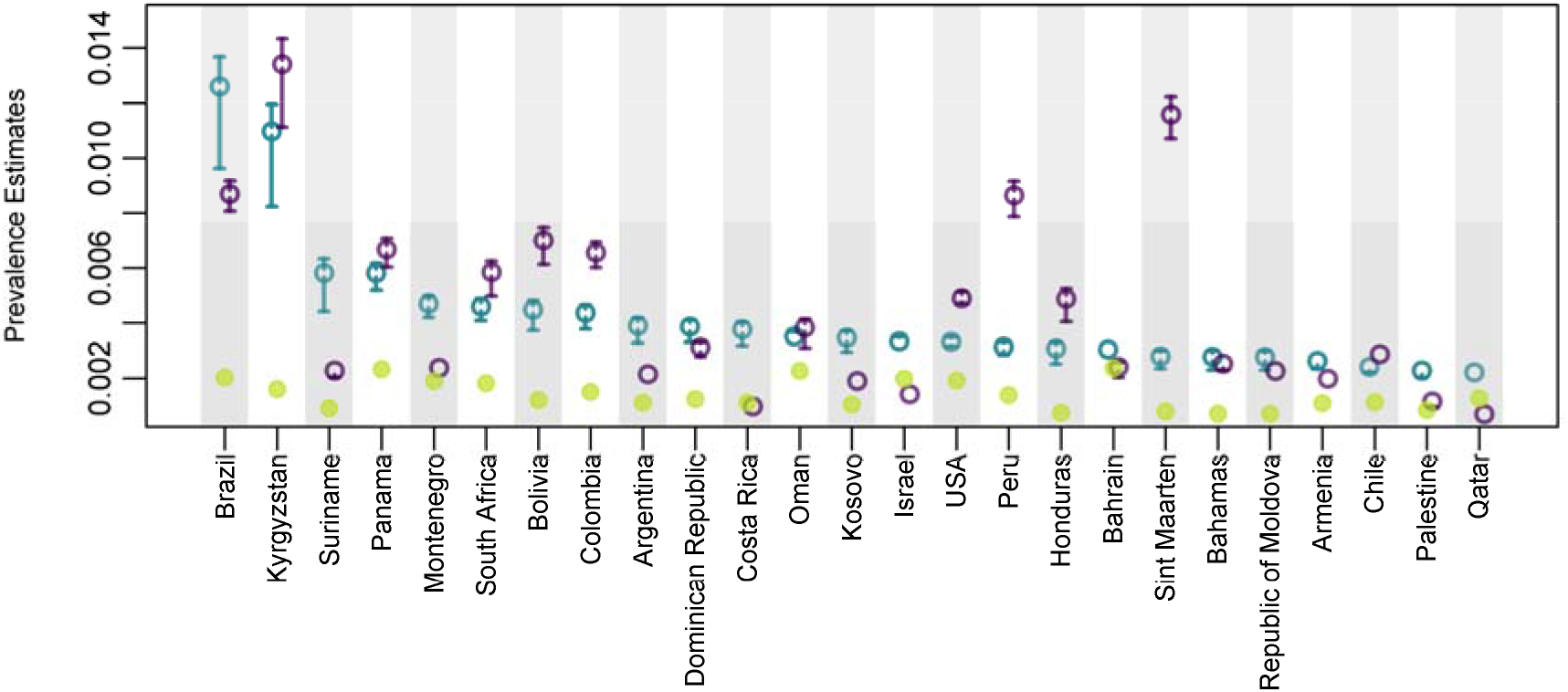
A comparison of the different stages used to produce the prevalence estimates in the countries with the top 25 prevalence. The raw prevalence (green circles) is adjusted to allow for the age distribution within the population (which impacts on the case fatality ratios, CFR and asymptomatic rates, ASR, purple circles). A country-specific underreporting factor after recalibrating for testing is then applied to create the final prevalence estimates (teal circles). Error bars show 95% confidence intervals.

Russell et al. have used a similar approach to estimating prevalence by adjusting for CFR (Russell *et al*., 2020), however they use a constant estimate of 1.4% for all countries. Our CFR estimates varied between 0.21% in Uganda to 2.2% in Japan. CFR have been shown to vary by age group (Verity et al. (2020) and Riou, Hauser, Counotte, and Althaus (2020)) and is a limitation that Russell *et al*. note.

We compare prevalence estimates between this model and the model proposed by Russell *et al*. for the 117 countries for which we could obtain estimates **(Error! Reference source not found.)-** The date for these prevalence estimates is the 12^th^ June 2020 which is the most recent update the authors have produced. In general, our estimates were more precise than the Russell *et al*. model, consistently producing smaller confidence intervals. Our model tended to produce lower prevalence estimates (82 of the 117 estimates, 70.08%). This is unsurprising as Russell *et al*. use an incidence rate estimate as a proxy to prevalence for their estimations. In instances where our model estimates a higher prevalence than the Russell *et al*. model, it generally occurred in countries with very low prevalence, and therefore there was very little difference between the prevalence rates. This does however suggest that our model may tend to overestimate the prevalence when there are a very small number of cases. Of the 117 estimates, 81 (69.23%) showed some degree of agreement however, with our estimates falling within the estimated 95% Credible Intervals of the Russell *et al*. model. In 41 countries, our prevalence estimates were significantly lower than the Russell *et al*. model, the majority of these countries tended to be ones with a high number of cases such as the US, Belgium, Sweden, Italy and the United Kingdom. The comparison within the UK is of particular interest as we estimate a prevalence of 0.046% (95%CI 0.043% - 0.048%) compared to the Russel *et al*. methodology which estimates 0.45% (95% CI 0.24% - 0.92%) which is ten times higher. The Office of National Statistics Infection pilot estimate for the period of 31^st^ May and 13^th^ June estimated a daily average prevalence in England to be 0.06% (95% CI 0.02% - 0.13%, (ONS, 2020a)). This would imply that our results may provide a more accurate estimate of prevalence, particularly in countries with a higher prevalence.

## Discussion

We have created an improved methodology to estimate the prevalence of a public health disease, and calculated the prevalence for all countries for COVID-19 disease where data for tests were available. The methods indicate that the prevalence of SARS-CoV-2 on 30^th^ July 2020 was highest in Brazil and Kyrgyzstan which correlates with the recent upsurge in cases in those countries. For example, there were less than 10,000 cases in Kyrgyzstan on June 30^th^ 2020 compared to 35,000 on July 30^th^ 2020. We have compared our model against other estimates of prevalence which are based on wide-scale testing (regardless of symptoms) for validation purposes, such as the ONS survey in the UK (ONS, 2020b). The most recent ONS survey results, for 20^th^-26^th^ July 2020, produce a weekly estimate of 0.07% (95% CI: 0.04% - 0.10%) prevalence in England. When we use our method for data up to 21^st^ July 2020, our estimate for the UK is 0.02% (95% CI 0.01 - 0.02). While our method produces a lower estimate, this is expected given we are estimating a date-specific prevalence rather than a weekly prevalence. Furthermore, our prevalence estimates are likely to be lower than ONS daily estimates in England as Northern Ireland and Scotland have lower case rates than England and Wales (Gov.uk, 2020). Further validation against other countries would allow better comparison between our model and true prevalence.

When we consider country prevalence and US state prevalence concurrently, Louisiana, Florida and Mississippi are the 3^rd^ to 5^th^ highest prevalence regions worldwide, even though when considered at a country level, the USA is only 15^th^ highest in the world. In fact, of the top 25 regions (countries or states), 15 of them are US states. This indicates the scale of the problem in some US states and across the country as a whole. In comparison, the majority of countries in Europe (37 out of the 46 which have testing data) are on a similar scale to the lowest prevalence US states (in the range 0 - 0.001).

The methodology incorporates disease-specific aspects, such as the length of time infected, age-dependent CFR and asymptomatic proportion, as well as a country’s pandemic response, by using a proxy of testing data. Both of these aspects are crucial to estimating effectively the prevalence in a country as shown by the impact they have on the estimates in some countries (Figure 4). By incorporating disease-specific considerations, the prevalence is able to more accurately represent the effect of the virus on the population, thereby turning this generic methodology into a specific one for SARS-CoV-2. The pandemic response is included due to the fact that not all countries are responding equally, with some countries testing many more individuals per population than others, and hence the number of reported cases will be more accurate. Similarly, some countries have better health systems, which will affect the likelihood of death given infection. The testing data is used as a proxy to represent all these pandemic response variables as it is significantly correlated with the difference between expected cases and reported cases. Without it, estimates for prevalence are generally much higher and less precise, as seen in the estimates produced by Russell *et al*. (2020). Our method combines country’s testing data by either number of tests performed or number of people tested which could lead to some bias as it is likely that the number of tests may be higher than the number of people tested. However, we found no statistical difference between these two numbers per population (p = 0.40).

Our improved methodology for estimating SARS-CoV-2 prevalence relies on widely available data for most countries around the world and hence is an easily applicable method for calculating prevalence quickly and timely. This allows for a horizon-scanning approach, to determine which countries are increasing or decreasing in prevalence, and therefore policy or aid can be changed accordingly. Furthermore, this methodology for SARS-CoV-2 prevalence is appropriate for similar public-health diseases that rely on testing of the population in order to determine the scale of the outbreak, especially those that may have a significant asymptomatic proportion, and hence reported cases are likely to be an underestimate. Only by having as much knowledge as possible will it be conceivable to reduce the spread of SARS-CoV-2 and hence this methodology to estimate prevalence, calculated on a regular basis, may aid countries in their response to this pandemic and any in the future.

## Data Availability

Data on the numbers of cases and deaths and number of tests for countries and territories were collated and provided by Public Health England (PHE) on the 30th of July 2020, based on either country specific public updates or data collated by the World Health Organisation. Population estimates were extracted from the Our World in Data website. Data for US states were also provided by PHE, collected from the Covidtracking.com website.

https://www.who.int/emergencies/diseases/novel-coronavirus-2019

https://ourworldindata.org/coronavirus-source-data

https://covidtracking.com/data

## Acknowledgements

We would like to thank Matthew Coleman (APHA) for review of the model and code and two anonymous reviewers (organised through the Rapid Assistance in Modelling the Pandemic (RAMP) group) for reviewing the model. This project was funded by Public Health England.

